# Assessment of multiplex digital droplet RT-PCR as an accurate diagnosis tool for SARS-CoV-2 detection in nasopharyngeal swabs and saliva samples

**DOI:** 10.1101/2020.08.02.20166694

**Authors:** Kévin Cassinari, Elodie Alessandri-Gradt, Pascal Chambon, Françoise Charbonnier, Ségolène Gracias, Ludivine Beaussire, Kevin Alexandre, Nasrin Sarafan-Vasseur, Claude Houdayer, Manuel Etienne, François Caron, Jean Christophe Plantier, Thierry Frebourg

## Abstract

RT-qPCR on nasopharyngeal swabs is currently the reference COVID-19 diagnosis method. We developed a multiplex RT-ddPCR assay, targeting six SARS-CoV-2 genomic regions, and evaluated it on nasopharyngeal swabs and saliva samples collected from 130 COVID-19 positive or negative ambulatory individuals, who presented symptoms suggestive of mild or moderate Sars-CoV2 infection. The 6-plex RT-ddPCR assay was shown to have 100% sensitivity on nasopharyngeal swabs and a higher sensibility than RT-qPCR on saliva (85% versus 62%). Saliva samples from 2 individuals with negative results on nasopharyngeal swabs were found to be positive. These results show that multiplex RT-ddPCR should represent an alternative and complementary tool for the diagnosis of COVID-19, in particular to control RT-qPCR ambiguous results, and its application to saliva an appropriate strategy for repetitive sampling and testing individuals for whom nasopharyngeal swabbing is not possible.

## INTRODUCTION

The reference biological method for the diagnosis of the new infectious Coronavirus disease 2019 (COVID-19), related to the severe acute respiratory syndrome coronavirus 2 (SARS-CoV-2), is the detection in the nasopharyngeal tract of the viral genome, using reverse transcription-quantitative PCR (RT-qPCR). Nasopharyngeal sampling is uncomfortable and the quality of sampling impacts the sensitivity of RT-qPCR (1). RT-qPCR has revealed highly variable viral loads among COVID-19 patients and, in a same patient, according to the time of sampling (2). Numerous RT-qPCR tests, targeting different regions of the viral genome, have been recently developed worldwide. Digital droplet PCR (ddPCR) represents an attractive alternative to qPCR. In ddPCR, the sample is separated in thousands of reactors and positive reactions are detected either with an intercalating agent or with hydrolysis probes (3). Therefore, this method enables the absolute quantification of nucleotide sequences by reducing the quantification of a target sequence to the enumeration of series of positive and negative end-point PCR reactions (4). ddPCR exhibits a higher analytical sensitivity and better reproducibility than qPCR, as shown by different applications in genetic (5) and viral diseases (6). Three studies, based on a 2-plex assay targeting two viral genomic segments, have already highlighted the potential of ddPCR for SARS-CoV-2 detection (7–9). One other advantage of ddPCR, as compared to qPCR, is the facility of multiplexing and this advantage has recently been illustrated for the detection of seasonal influenza virus (10). Moreover, some studies have recently evaluated the interest of RT-qPCR performed on saliva for SARS-CoV-2 detection. These studies have shown that this strategy, as compared to RT-qPCR on nasopharyngeal swabs, has a highly variable sensitivity (30.7%-100%), depending in particular on the mode and conditions of saliva collection (11, 12).

In this study, we developed and validated a COVID-19 multiplex RT-ddPCR assay, including six probe-primer sets already validated in qPCR assays and then evaluated the performances of the assay for the detection of SARS-CoV-2 in nasopharyngeal and saliva samples collected in a cohort of patients.

## PATIENTS AND METHODS

### Patients

For the validation step of RT-ddPCR, we selected a series of nasopharyngeal swabs with a low viral load, defined on the basis of a Cycle Threshold (CT) >30 in RT-qPCR. For the prospective phase of the study, biological samples were collected from patients presenting at the COVID-19 consultation of Rouen University Hospital, during the first epidemic peak in our area (from April to May 2020). All patients were ambulatory and presented symptoms suggestive of mild or moderate Sars-CoV2 infection. All patients had a deep nasopharyngeal swabbing (Sigma Virocult® system -MWE, Corsham,UK), and then were asked to drool around 2 mL of saliva into a sterile 50 mL Falcon plastic tube (Thermo Fisher Scientific, Illkirch, France), after they gave informed consent. Samples were transferred within 2 h to the laboratory of Virology and then were frozen before subsequent RNA extraction. For all patients with positive RT-qPCR in nasopharyngeal samples, the corresponding saliva was then analyzed by RT-qPCR and RT-ddPCR. Saliva was also analyzed by RT-qPCR and RT-ddPCR in a subset of subjects with negative nasopharyngeal swabs. The protocol was approved by the institutional ethics committee (2020T3-12_RIPH3 HPS_2020-A00920-39).

### RT-qPCR

After viral inactivation, two qualitative methods of RT-qPCR were used by the Virology laboratory of Rouen University Hospital for routine diagnosis, depending on the supply stock: (i) an automated method, using Abbott RealTime SARS-CoV-2 EUA test (Abbott Park, IL, USA), performed on 500 µl of nasopharyngeal samples and (ii) RNA extraction from 200 µl of sample (nasopharyngeal swab or saliva), performed using EZ1 DSP virus kit (Qiagen, Hilden, Germany) and EZ1 Advanced XL machine, then RT-qPCR on 10µl of extracted RNA, using RealStar® SARS-CoV-2 RT-PCR Kit 1.0 (Altona Diagnostics, Hamburg, Germany) and performed on a CFX96™ Real-Time PCR Detection System (BioRad, Californie, USA).

### RT-ddPCR

RT-ddPCR assays were performed using the One-Step RT-ddPCR Advanced Kit for Probes (Bio-Rad Laboratories, Hercules, CA, USA) and the QX200 ddPCR platform (Biorad). Firstly, a 2-plex RT-ddPCR assay was developed on the basis of the French national reference centre COVID-19 RT-qPCR protocol, which targets two regions of the RdRp gene: nCoV_IP2 and nCoV_IP4. Secondly, a 6-plex RT-ddPCR assay including 4 additional targets was developed (Supplementary Table 1). These additional targets were selected among primers and probes referenced in the Open COVID-19 Testing Project (https://covidtestingproject.org/), on the basis of the amplicon size (<120 bp). The specificity of each primer and probe was checked using the BLASTN program (https://blast.ncbi.nlm.nih.gov/) across a bank of 2045 viral genomic sequences. All hydrolysis probes were designed with a FAM or HEX fluorophore and quenchers optimized for ddPCR (Iowa Black quencher and an internal ZEN quencher, IDT DNA).

Briefly, 9.5 μL of extracted RNA was diluted in a 22 μL final reaction volume containing 5.5 μL of One Step SuperMix (ddPCR supermix for Probes no dUTP, Bio-Rad), 2.2 μL of Reverse Transcriptase, 1.1 μL of 300mM DTT and 3 μL of primers and probes mix (final probe concentration: 250 nM each, final primer concentration: 750 nM each). Then, each sample was partitioned into 13000 to 20000 droplets using the QX200 ddPCR Droplet Generator System (Bio-Rad). PCR amplification was then performed on a T1000 thermal cycler (Bio-Rad). This protocol included an initial retro-transcription step (60 min, 50°C, and 10 min, 95°C) followed by 40 cycles of cDNA amplification, each cycle including a denaturation step (95°C for 30 sec) and a step of annealing and extension at (58°C for 1 min). A final denaturation step was realized at 98°C for 10 min. The droplet reading and quantification were performed using the QX200 droplet digital reader and data analysis was performed using the 2D module of the QuantaSoft-Pro software (Bio-Rad). Limit of blank (LOB) and limit of detection (LOD) of the assay were determined according to published guidelines (13, 14).

## RESULTS

### Development and validation of the multiplex RT-ddPCR assay

For the development of the RT-ddPCR assay, we compared on previously collected nasopharyngeal samples the sensitivity of a 2-plex RT-ddPCR assay targeting nCoV_IP2 and nCoV_IP4 (respectively detected in the HEX and FAM channel), to that of a 6-plex RT-ddPCR assay (Fig. 1a), targeting three genomic regions detected in the FAM channel (CN-CDC-1, CN-CDC-2, nCoV-IP4) and three detected in the HEX channel (nCoV_N1, nCoV_IP2, RdRp_Sars_r). As expected, for a given viral load, a higher number of positive droplets was detected with the 6-plex assay, as compared to the 2-plex assay (Fig. 1b). After calculation of the LOD (Supplementary Tables 2 and 3), analysis of serial dilutions of a highly positive sample showed the higher sensitivity of the 6-plex RT-ddPCR assay (Supplementary Table 4). We validated the 6-plex RT-ddPCR assay by analysing 50 low positive nasopharyngeal samples, as determined by RT-qPCR. All these samples were unambiguously detected positive by the 6-plex RT-ddPCR assay (Supplementary Table 5).

**FIG 1:**
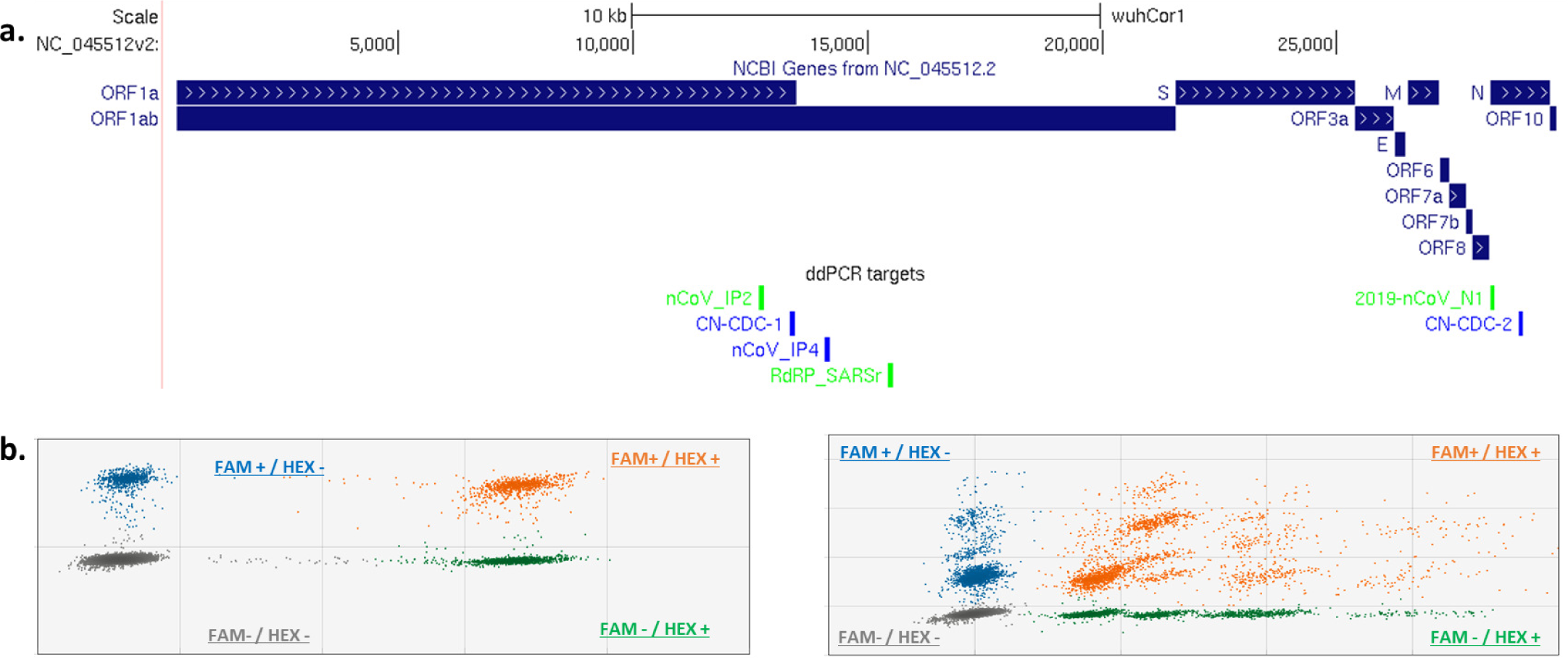
Presentation of the 6-plex RT-ddPCR for SARS-CoV-2 detection. **(a)** Visualization, using the UCSC Genome Browser (Santa Cruz University, https://genome.ucsc.edu) of the SARS-CoV-2 genome (NC_045512v2). Top panel: in blue, list and location of genes (NC_045512.2). Bottom panel: custom track, indicating the genomic location of each targeted integrated in the 6-plex RT-ddPCR (blue: FAM labelled; green: HEX labelled). (**b)** Representative examples of positive 2-plex (left panel) and 6-plex (right panels) RT-ddPCR assays performed on a RT-qPCR positive nasopharyngeal swab. All spots, except the grey ones, represent positive droplets containing viral genomic material. The 2-plex RT-ddPCR assay targets nCoV_IP2 and nCoV_IP4 located within the RNA-dependent RNA polymerase (RdRp) gene. Blue droplets (FAM fluorescence): positive for the IP4 target; green droplets (HEX fluorescence): positive for the IP2 target; orange droplets (FAM and HEX fluorescence): positive for both IP4 and IP2 targets; grey droplets (no fluorescence): negative. The 6-plex RT-ddPCR assay targets six regions of the viral genome: nCoV_IP2, nCoV_IP4, nCoV_CDC-1 and RdRp_SARSr located within the RdRp gene, N_CoV_N1 and CN_CDC-2 within the Neuramidase gene. Blue droplets (FAM fluorescence): positive for the N_CoV_IP4, CN-CDC-1 and/or CN-CDC-2 targets; green droplets (HEX fluorescence): positive for the N_CoV_IP2, RdRP_SARsR and/or nCoV_N1 targets; orange droplets (FAM and HEX fluorescence): positive for at least one FAM-labelled target and one HEX-labelled target, and grey droplets (no fluorescence): no target. According to the total of FAM and HEX droplets, the results were estimated in the 2-plex and 6-plex RT-ddPCR assays to 7234 and 18820 copies per reaction, respectively.

### Prospective analysis of nasopharyngeal swabs and saliva samples using the 6-plex RT-ddPCR assay

We then prospectively collected nasopharyngeal swabs and saliva samples from 130 patients and 14 were found to have a positive RT-qPCR test on nasopharyngeal swabs. Among the 14 corresponding saliva samples, one was excluded because of an insufficient volume. RT-qPCR analysis of these 13 saliva samples yielded 8 positive and 5 negative results, corresponding to a sensitivity of 62% (Supplementary Table 6). The 6-plex RT-ddPCR assay was also positive on the 14 nasopharyngeal swabs. The 6-plex RT-ddPCR assay performed on the 13 saliva samples retrieved 11 positive (including one sample positive and negative in the FAM and HEX channels, respectively) and 2 negative results, indicating a sensitivity of 85% (Supplementary Table 6). The mean ratio of the SARS-CoV-2 load between nasopharyngeal swabs and saliva was estimated, according to RT-ddPCR, to 457 with a very large inter-individual variation (Supplementary Table 6).

The 6-plex RT-ddPCR assay was also performed on a subset of 18 saliva collected from 116 patients with a negative RT-qPCR test on nasopharyngeal swab. For these 18 patients, RT-ddPCR was also negative on nasopharyngeal swabs (Supplementary Table 7). Interestingly, one of the saliva sample (ID: 007) was found positive by RT-ddPCR, this result was confirmed by a second RT-ddPCR analysis and by RT-qPCR which was subsequently performed. For another saliva sample (ID: 017), we obtained values just above the LOD (Supplementary Table 7).

## DISCUSSION

We show in this study the interest of multiplex RT-ddPCR for the diagnosis of COVID-19. The global sensibility of COVID-19 molecular diagnostic methods depends on several factors including the quality of sampling, the integrity of viral RNA, the efficiency of RNA extraction and PCR. As RNA degradation or imperfect retro-transcription of RNA templates may hamper the detection of SARS-CoV-2, especially in samples with a very low viral load, increasing the number of viral targets within the same assay should improve the sensibility of the assay and this is confirmed by our results showing the higher sensibility of the 6-plex RT-ddPCR assay, as compared to the 2-plex PCR assay. Target multiplexing is much easier to be performed in RT-ddPCR than in RT-qPCR. Indeed, ddPCR relies on a final PCR point and does not require, in contrast to RT-qPCR, to optimize PCR conditions for each target. Although target multiplexing has been shown to increase the background noise because of non-specific probes hydrolysis (10), our results show that this does not represent a technical limit of the COVID-19 multiplex RT-dPCR assay.

The results that we obtained on nasopharyngeal swabs with a low viral load, as estimated by CT for RT-qPCR, show that RT-ddPCR could be used either as a complementary method to re-analyze samples yielding ambiguous results in RT-qPCR or as an alternative method requiring different reagents and platforms. The analysis of a limited series of saliva samples show that this assay can also be applied to saliva. As previously reported, we confirm that the sensibility of COVID-19 molecular assays performed in saliva is significantly weaker than in nasopharyngeal swabs, probably because of a viral load in average 400 times lower. We show a slightly higher sensibility of the multiplex RT-ddPCR assay (85%), as compared to RT-qPCR (62%) for saliva analyses. It should be highlighted that saliva samples were collected in this study without any specific conditions, while it has been shown that collection of saliva after overnight fasting result in a higher RNA concentration [15]. Therefore, we think that it might be possible to increase the sensibility of the multiplex RT-ddPCR assay on saliva with specific conditions of saliva sampling.

One advantage of the SARS-CoV-2 RT-ddPCR assay is its potential to be optimized: (i) Indeed, its sensitivity can probably be increased by the addition of other SARS-CoV-2 targets regularly spaced across the viral genome and corresponding to short amplicons (around 70bp), in order to prevent the co-encapsulation of several viral genomic targets within the same droplet. Multiplexing of viral targets is also of interest in the perspective of the mutability of the SARS-CoV-2 genome, which might result in false negative results yielded by molecular assays restricted to a single genomic region; (ii) The assay can also be optimized by the integration of a human gene which would allow to evaluate not only the cellularity of the sample and thus the quality of the sampling, but also the quality of RNA extraction and PCR.

As saliva sampling is a non-invasive collection procedure, it represents an appropriate strategy to test repeatedly individuals (e.g. in nursing homes), to test individuals for whom nasopharyngeal swabs are contraindicated or to test large populations suspected to present with a high viral load. In this context, we think that the multiplex RT-ddPCR, which sensitivity may probably be optimized, should be of interest. Our study also illustrates one of the limits of nasopharyngeal swabs. Among 18 individuals with a molecular test negative on nasopharyngeal swabs, the saliva sample was found to be clearly positive in one patient. It reinforces the hypothesis that some of the false negative RT-qPCR results could be related to a lower cellularity of nasopharyngeal swabs, due to suboptimal sampling (1). This idea is supported by the observation that, in a few samples, RT-ddPCR detected a higher viral concentration in saliva than in nasopharyngeal swab. Therefore, saliva sampling may also be considered as supplementary sample in patients with negative tests on nasopharyngeal swabs but with symptoms strongly suggestive of COVID-19.

## Data Availability

Anonymized data relevant to this study and not included here will be made available upon reasonable request.

## Acknowledgements

We thank Mathieu Castelain, Charlène Martin for logistic support and Anne-Claire Richard, Fabienne De Oliveira for technical assistance.

